# GlioMODA: Robust Glioma Segmentation in Clinical Routine

**DOI:** 10.1101/2025.11.12.25339968

**Authors:** Julian Canisius, Josef Buchner, Marcel Rosier, Michael Griessmair, Jan Peeken, Jan S Kirschke, Marie Piraud, Spyridon Bakas, Bjoern Menze, Benedikt Wiestler, Florian Kofler

**Author notes:** Corresponding author: Julian Canisius, MD, Institute of Neuroradiology, Klinikum rechts der Isar, Technical University of Munich (TUM), Ismaninger Str. 22, 81675 Munich, Germany. Equal contribution: Julian Canisius and Josef Buchner contributed equally as co-first authors. Shared senior authorship: Benedikt Wiestler and Florian Kofler contributed equally as co-senior authors.

## Abstract

**Background:** Precise glioma segmentation in MRI is essential for accurate diagnosis, optimal treatment planning, and advancing clinical research. However, most deep learning approaches require complete, standardized MRI protocols that are frequently unavailable in routine clinical practice. This study presents and evaluates GlioMODA, a robust deep learning framework designed for automated glioma segmentation that delivers consistent high performance across varied and incomplete MRI protocols.

**Methods:** GlioMODA was trained and validated on the BraTS 2021 dataset (1,251 training, 219 testing cases), systematically assessing performance across eleven clinically relevant MRI protocol combinations. Segmentation accuracy was evaluated using Dice similarity coefficients (DSC) and panoptic quality metrics. Volumetric accuracy was benchmarked against manual ground truth, and statistical significance was established via Wilcoxon signed-rank tests with Benjamini–Yekutieli correction.

**Results:** GlioMODA demonstrated state-of-the-art segmentation accuracy across tumor subregions, maintaining robust performance with incomplete or heterogeneous MRI protocols. Protocols including both T1-weighted contrast-enhanced and T2-FLAIR sequences yielded volumetric differences versus manual ground truth that were not statistically significant for enhancing tumor (ET: median difference 55 mm³, p = 0.157) and whole tumor (WT: median difference –7 mm³, p = 1.0), and exhibited median DSC differences close to zero relative to the four-sequence reference protocol. Omitting either sequence led to substantial and significant volumetric errors.

**Conclusions:** GlioMODA facilitates reliable, automated glioma segmentation using a streamlined two-sequence protocol (T1-contrast + T2-FLAIR), supporting clinical workflow optimization and broader implementation of quantitative volumetry compatible with RANO 2.0 criteria. GlioMODA is published as an open-source, easy-to-use Python package at https://github.com/BrainLesion/GlioMODA/.

**Key Points:**

- T1-CE + T2-FLAIR maintains enhancing and whole tumor segmentation comparable to four-sequence MRI.
- Consistent volumes with T1-CE + T2-FLAIR support reliable RANO 2.0 assessment.
- Open-source GlioMODA (models + code) supports rapid integration.

**Importance of the Study:** Automated glioma segmentation is limited in practice by incomplete or heterogeneous MRI protocols. GlioMODA directly addresses this barrier by delivering consistent accuracy across 11 clinically relevant sequence combinations and identifying a streamlined protocol (T1-contrast and T2-FLAIR) whose enhancing- and whole-tumor volumes are not statistically different from expert reference. This enables shorter scans and reproducible volumetry compatible with RANO 2.0, facilitating reliable response assessment in trials and routine care. By releasing trained models and code as an easy-to-use open-source package, this work enables external validation and integration into neuro-oncology workflows.

## Introduction

Gliomas are the most common primary brain tumors in adults, with glioblastoma multiforme (GBM) representing the most aggressive subtype, characterized by imaging heterogeneity, diffuse infiltration, rapid progression, and poor prognosis (1,2). Magnetic resonance imaging (MRI) is the diagnostic standard for GBM, enabling visualization of key tumor subregions essential for treatment planning, prognosis assessment, and monitoring therapeutic response (3,4). These subregions include enhancing tumor (ET), tumor core (TC), and whole tumor (WT), with accurate segmentation central for clinical decision-making. The tumor core, including the enhancing tumor, is essential for defining the gross tumor volume (GTV) in postoperative radiotherapy following the Stupp protocol, while edema segmentation is vital for assessing the clinical target volume (CTV), which encompasses infiltrative tumor areas, including non-enhancing, FLAIR-hyperintense regions (5,6). Manual delineation of these subregions remains challenging due to the complex and heterogeneous structure of gliomas, making the process time-consuming, labor-intensive, and subject to substantial inter- and intra-rater variability in both clinical practice and research (7,8).

In recent years, automated segmentation algorithms based on convolutional neural networks (CNNs), particularly U-Net variants, have achieved expert-level accuracy and reduced clinician workload (4). These algorithms substantially reduce the time required for manual segmentation while achieving accuracy comparable to expert reference segmentations, often within interrater variability. In addition, automated segmentation has demonstrated prognostic relevance, with volumetric measurements showing significant associations with survival outcomes in glioma patients (9). Furthermore, automated approaches help standardize response assessment across institutions, reducing inter-observer variability that affects treatment evaluation (10). However, translation to routine care requires robust performance under incomplete or heterogeneous MRI protocols, compatibility with contemporary response frameworks that accommodate quantitative volumetry (e.g., RANO 2.0) under standardized imaging conditions (11).

Yet most state-of-the-art approaches, such as those evaluated in the Brain Tumor Segmentation (BraTS) challenge (3,12), assume complete, standardized four-sequence MRI—T1-weighted native (T1n), T1-weighted contrast-enhanced (T1c; often referred to as T1-CE), T2-weighted (T2w), and T2-FLAIR (T2f) (13). Hereafter, the abbreviations T1n, T1c, T2w, and T2f are used throughout. A standardized brain tumor imaging protocol for glioblastoma including these four MRI sequences averages a scan time of approximately 21 minutes; acquiring all four sequences increases scan time, cost, and patient burden; and protocol variability or incomplete data are common in routine care (14). The prevalence of this issue can be significant; for instance, a multicenter glioma dataset reported missing sequences exceeding 20% (15).

Recent advances in deep learning suggest that sequence requirements for high-quality glioma segmentation can be reduced without significantly compromising segmentation performance (16). Concordantly, Buchner et al. previously identified core MRI sequences for reliable automatic brain metastasis segmentation. Here, T1-CE alone yielded strong lesion segmentation, with T2-FLAIR necessary to capture surrounding FLAIR-hyperintense edema (17). More broadly, recent studies indicate that certain MRI modalities may be redundant for segmentation tasks, and that reduced-input models - using only the most informative sequences - can achieve segmentation performance comparable to full-input models, offering practical advantages in terms of efficiency and applicability. Shorter protocols prodvide clinical advantages, including lower motion risk (14), greater patient comfort, reduced costs, improved workflow efficiency (4), and minimized risks associated with contrast agents (18). They also lower the amount of data that needs to be processed, enabling faster pre-processing times and leaner models, which is advantageous for high-throughput and resource-limited environments.

Moreover, accurate delineation of these tumor subregions provides valuable information for downstream analysis with techniques such as radiomics or neural network-based feature extraction, supporting non-invasive characterization, molecular prediction, and outcome modeling in glioma and brain metastasis patients (19,20).

Despite these advances, there remains limited determination of which MRI sequence combinations are essential to maintain robust glioma segmentation across real-world scenarios. Addressing this gap is crucial to enable automated analysis when protocols are incomplete and to optimize clinical workflows (21), also aligning with contemporary response frameworks that increasingly accommodate quantitative volumetry (e.g., RANO 2.0) (11).

Therefore, this study systematically evaluates GlioMODA—a flexible deep learning framework for automated glioma segmentation—across all clinically relevant combinations of standard MRI sequences. Our aim is to identify core MRI sequences that enable accurate, efficient, and broadly applicable glioma segmentation, thereby supporting protocol reduction, improving clinical workflow, and expanding access to advanced image analysis in routine practice. By delineating sequence combinations that preserve clinical-grade accuracy while reducing acquisition time, this work aims to facilitate translation into routine neuro-oncology practice and contemporary quantitative response assessment, ultimately improving patient care.

## Methods

### Dataset

We used the BraTS preoperative glioma dataset from 2021 (22), comprising 1,251 training cases and 219 testing cases with histologically confirmed gliomas from multiple international institutions. Following standard practice in machine learning, all models were trained exclusively on the 1,251-case training cohort without using a separate validation set, while the 219-case cohort (originally employed as the “validation set” in the BraTS 2021 segmentation challenge) served as our independent test set and remained unseen throughout the training process. All cases included complete preoperative multiparametric MRI with standardized four-sequence protocols: T1-weighted native (T1n), T1-weighted contrast-enhanced (T1c), T2-weighted (T2w), and T2-FLAIR (T2f) images. Expert manual segmentations of central necrosis (NEC), contrast-enhancing tumor (CET), and surrounding edema (ED) were provided following rigorous quality control by board-certified neuroradiologists. The multicenter nature of this dataset supports generalizability across diverse clinical environments and imaging protocols. Detailed annotation protocols and data collection procedures are described in the original BraTS 2021 publication (22).

### MRI sequence combinations

To systematically evaluate the impact of protocol completeness on glioma segmentation, we analyzed 11 clinically relevant MRI sequence combinations (Table 1). These were selected based on the Brain Tumor Imaging Protocol (BTIP) guidelines for glioblastoma (13) and prior evidence of sequence utility in tumor subregion delineation (6,22).

**Table 1.**
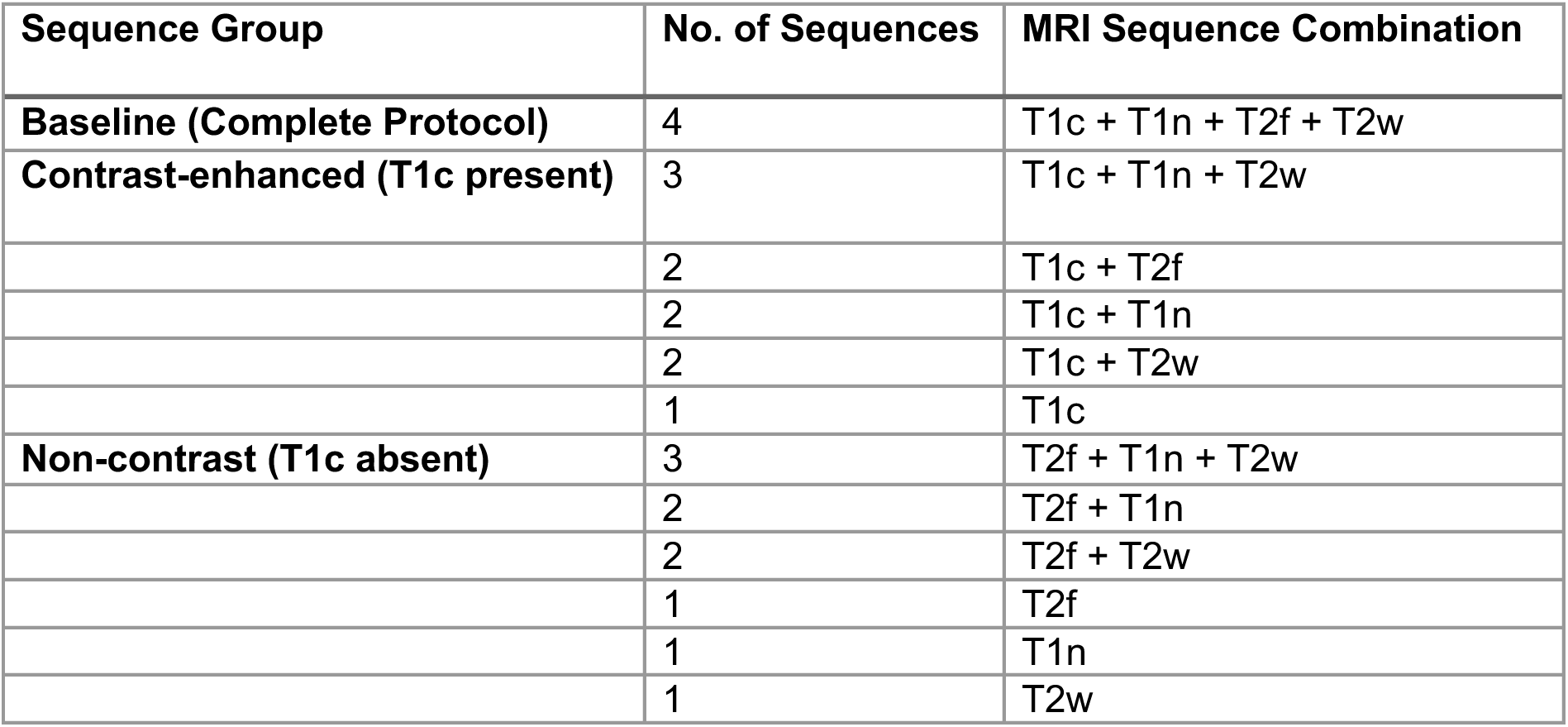
MRI sequence combinations. Sequence groups, number of sequences, and protocol compositions evaluated in this study. Abbreviations: T1c = T1-weighted contrast-enhanced; T1n = T1-weighted native; T2f = T2-FLAIR; T2w = T2-weighted.

| Sequence Group | No. of Sequences | MRI Sequence Combination |
| --- | --- | --- |
| **Baseline (Complete Protocol)** | 4 | T1c + T1n + T2f + T2w |
| **Contrast-enhanced (T1c present)** | 3 | T1c + T1n + T2w |
|  | 2 | T1c + T2f |
|  | 2 | T1c + T1n |
|  | 2 | T1c + T2w |
|  | 1 | T1c |
| **Non-contrast (T1c absent)** | 3 | T2f + T1n + T2w |
|  | 2 | T2f + T1n |
|  | 2 | T2f + T2w |
|  | 1 | T2f |
|  | 1 | T1n |
|  | 1 | T2w |

### Neural Network

We used the nnU-Net framework (version 2.5.2) to train and test our neural networks (23). After analyzing the dataset fingerprint for each sequence combination, we trained a neural network according to the chosen experiment planner for a total of 1,000 epochs.

Although Isensee et al. recommend the residual-encoder U-Net presets cross-vaas the new default (24), we found no performance differences compared to the previous default experiment planner, despite substantially increasing the training duration on our hardware. Therefore, we chose to use the default planner. We used the 3D-fullres configuration.

Furthermore, cross-validation was not performed; instead, we trained on all available training data. Note that the 219 validation patients were never used during training, but only for final model testing.

Following the BraTS challenge evaluation protocol, we did not evaluate the individual labels but rather combined them into clinically significant subregions: enhancing tumor (ET), tumor core (TC), and whole tumor (WT). To reflect this during training, we used the region-based training that is already integrated into the nnU-Net.

All training runs were conducted on Nvidia RTX 6000/8000 GPUs using CUDA version 12.2. Each training run took around 20 hours.

### Metrics

Segmentation performance was primarily evaluated using the Dice similarity coefficient (DSC) (25), quantifying spatial overlap between predicted and reference segmentations for the enhancing tumor (ET), tumor core (TC), and whole tumor (WT) subregions. For comprehensive assessment, we also report panoptic quality (PQ), combining segmentation quality (SQ) and recognition quality (RQ) to reflect both boundary accuracy and lesion detection (Figure 2) (26). All metrics were calculated with the Python package panoptica (27).

For Figures 1 and 3, segmentation performance for each sequence combination was compared to the four-sequence baseline (T1c+T2f+T1n+T2w), with DSC differences (Figure 1) and corresponding Wilcoxon signed-rank tests (28) to assess statistical significance. The resulting p-values were adjusted for multiple comparisons using the Benjamini–Yekutieli procedure (29) and visualized as heatmaps (Figure 3).

**Figure 1.**
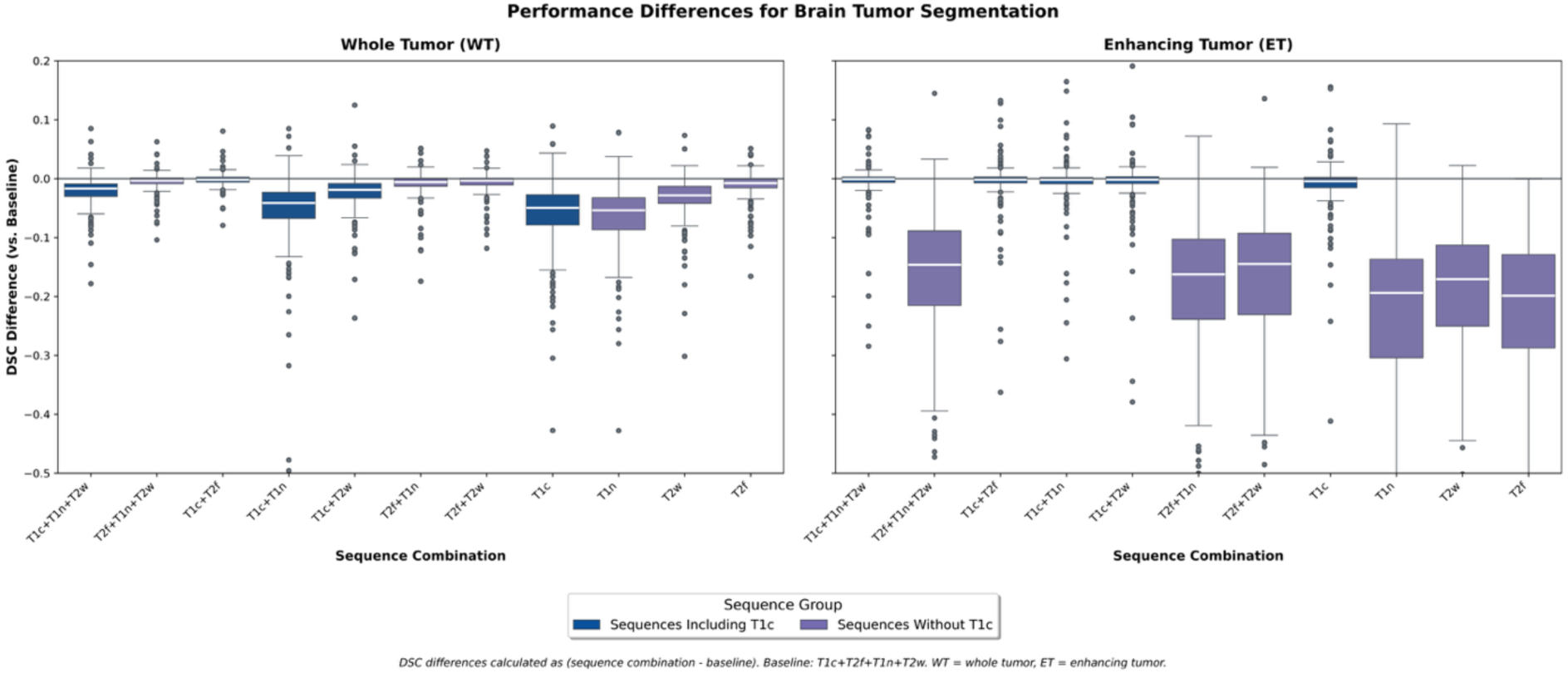
Dice similarity coefficient differences relative to the four-sequence baseline. Box plots illustrate Dice similarity coefficient (DSC) differences from the four-sequence baseline protocol (T1c+T2f+T1n+T2w) for whole tumor (WT) and enhancing tumor (ET) segmentation. Boxes show the median (white line) and interquartile range (IQR); whiskers extend to 1.5× IQR, and points indicate outliers. Abbreviations: DSC, ET, WT, T1c, T1n, T2f, T2w.

**Figure 2.**
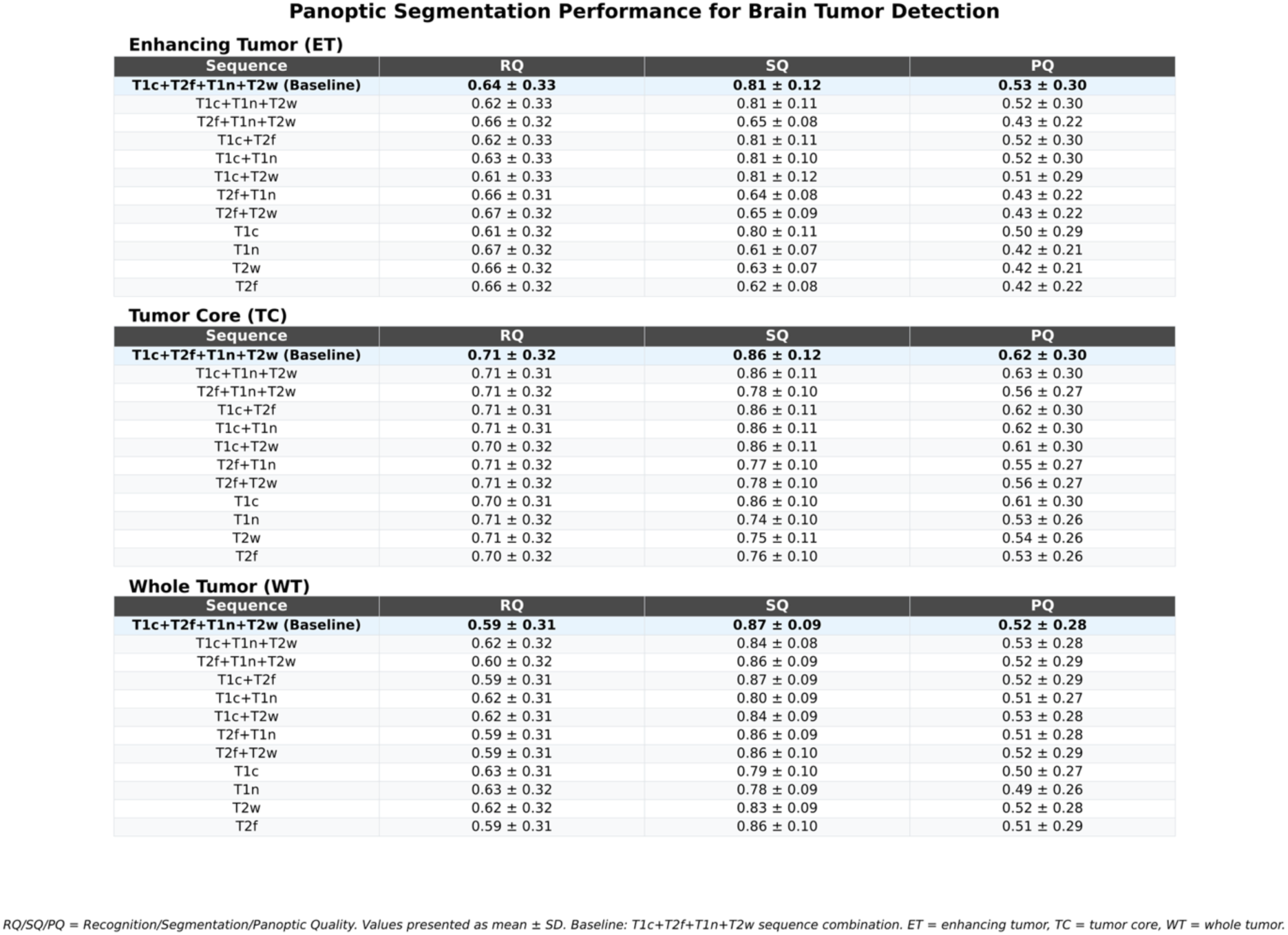
Panoptic assessment across MRI sequence combinations. Recognition quality (RQ), segmentation quality (SQ), and panoptic quality (PQ) are reported for ET, tumor core (TC), and WT across all protocol combinations; values are presented as mean ± SD. RQ reflects instance detection (F1-equivalent with an overlap threshold), SQ reflects boundary accuracy for true positives, and PQ combines both measures.

**Figure 3.**
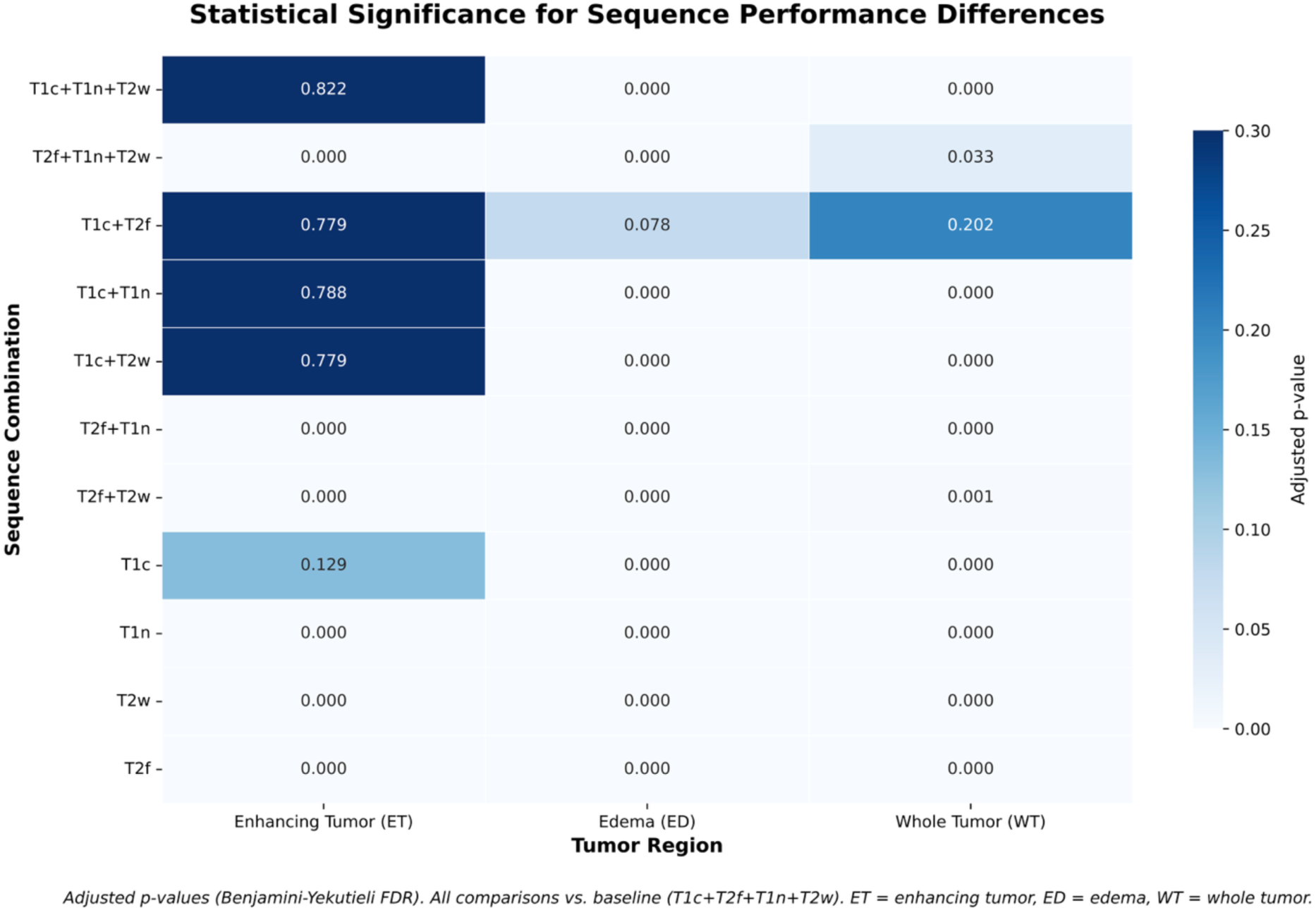
Statistical significance for sequence performance differences. The heatmap displays adjusted p-values from Wilcoxon signed-rank tests comparing each sequence combination with the four-sequence baseline across ET, edema (ED), and WT. P-values were adjusted using the Benjamini–Yekutieli procedure for multiple comparisons correction. Color intensity represents statistical significance levels, with darker shades indicating higher p-values (non-significant differences) and lighter shades showing lower p-values (significant differences).

For Figures 4 and 5, all protocol combinations—including the baseline—were compared directly to manual ground truth by assessing absolute volumetric errors (Figure 4). Statistical significance of these volumetric differences was evaluated using the same Wilcoxon signed-rank tests with Benjamini–Yekutieli correction, and results were visualized as heatmaps in parallel style and colormap as used for DSC analyses (Figure 5), allowing direct visual and statistical comparison.

**Figure 4.**
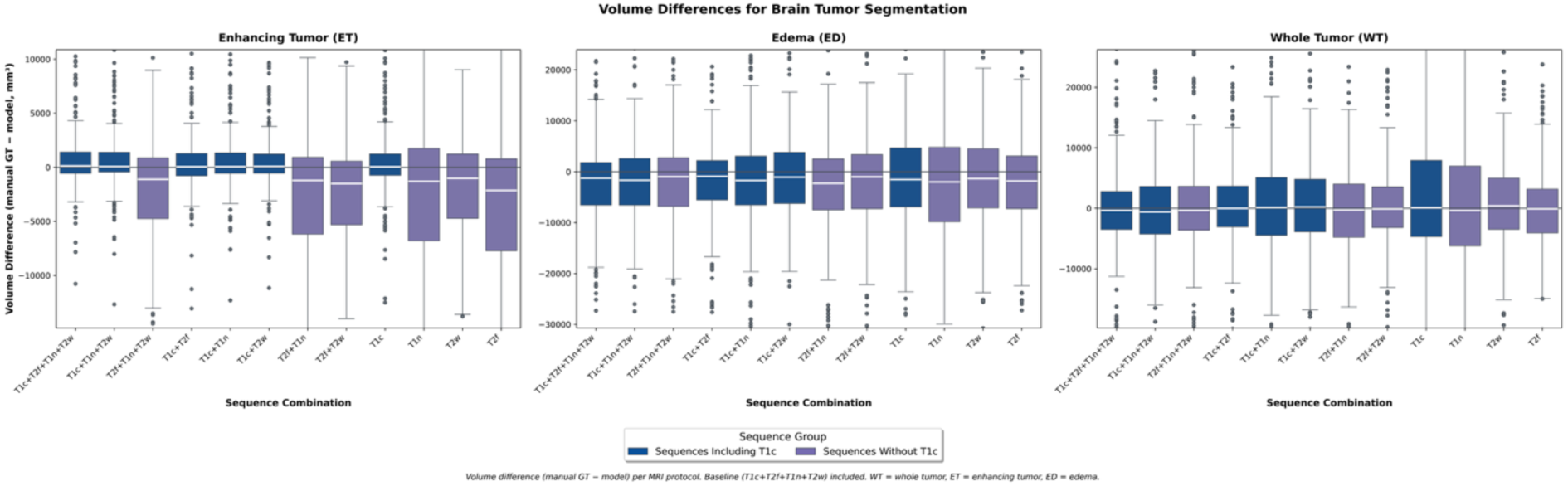
Volumetric accuracy across MRI sequence combinations. Box plots show absolute volume differences between model segmentation and manual ground truth (manual GT – model, mm³) for ET, ED, and WT across MRI sequence combinations. Negative values indicate over-segmentation and positive values indicate under-segmentation relative to ground truth. Boxes show the median (white line) and IQR; whiskers extend to 1.5× IQR. The baseline protocol is T1c+T2f+T1n+T2w.

**Figure 5.**
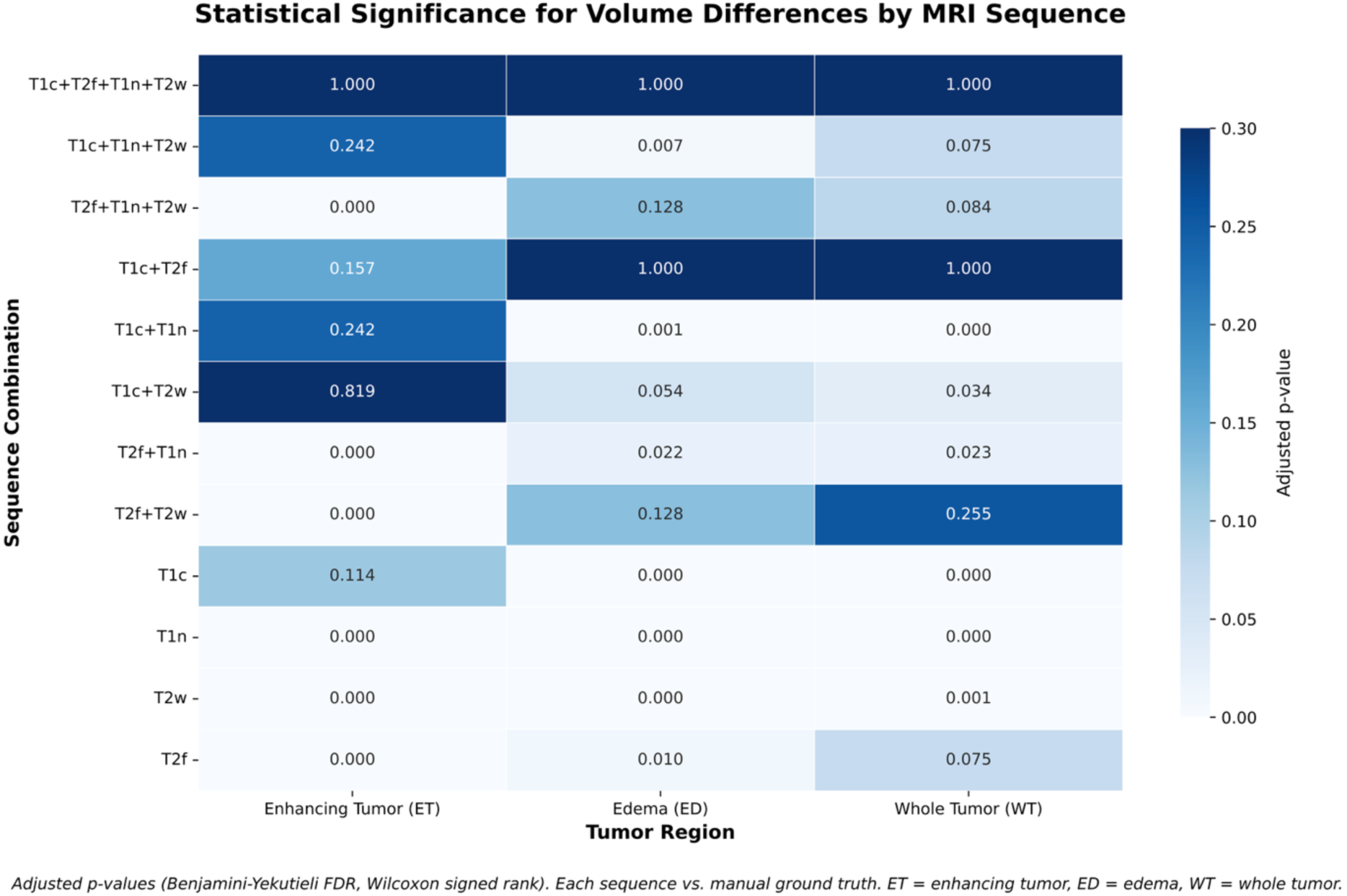
Statistical significance of volumetric differences across MRI sequence combinations. The heatmap shows Benjamini–Yekutieli–adjusted p-values (Wilcoxon signed-rank) for volumetric differences between model and manual ground truth across sequence combinations and tumor regions (ET, ED, WT). Color intensity corresponds to adjusted p-value, with lighter shades reflecting stronger statistical significance (p < 0.05).

All results are reported as median (interquartile range) across the test cohort. Statistical significance was defined as an adjusted p-value < 0.05.

Difference plots, box plots, and statistical significance heatmaps were generated using Matplotlib (30) and seaborn (31), with protocol-specific color schemes and consistent colormap use for both DSC and volumetric analyses to facilitate clear interpretation and comparability across all figures.

Reporting follows the CLEAR checklist for radiomics and AI imaging studies; the completed checklist is provided as Supplementary Material.

## Results

### Sequence Performance and Quality Assessment

To systematically evaluate MRI sequence combinations for glioblastoma segmentation, we compared all possible combinations to the four-sequence baseline (T1c+T2f+T1n+T2w) using DSC differences and comprehensive panoptic segmentation metrics (Figures 1–2).

For enhancing tumor (ET) segmentation, T1c-containing combinations demonstrated superior performance with DSC differences clustered tightly around zero. T1c alone achieved near-baseline performance with minimal deviation (median: –0.004, IQR: –0.015 to +0.002), while T1c combinations showed even tighter distributions: T1c+T2f (median: –0.002, IQR: – 0.007 to +0.003), T1c+T1n (median: –0.002, IQR: –0.009 to +0.002), and T1c+T2w (median: –0.002, IQR: –0.008 to +0.003). Panoptic segmentation analysis (Figure 2) confirmed these findings, with T1c alone achieving recognition, segmentation, and panoptic quality scores (0.61 ± 0.32, 0.80 ± 0.11, 0.50 ± 0.29) comparable to baseline (0.64 ± 0.33, 0.81 ± 0.12, 0.53 ± 0.30). All T1c combinations maintained consistently high segmentation quality scores of 0.80–0.81, matching or exceeding baseline performance. In contrast, combinations lacking T1c showed substantial performance degradation with DSC differences of –0.171 to –0.199 and markedly wider variability ranges. Segmentation quality scores dropped to 0.61–0.65 for non-T1c combinations, representing a clinically meaningful 0.16–0.20 decrease compared to baseline and confirming T1c’s fundamental importance for accurate ET boundary delineation.

For whole tumor (WT) segmentation, T2f-containing combinations provided optimal performance stability across all evaluated metrics. The T1c+T2f combination achieved near-baseline performance (median DSC difference: –0.002, IQR: –0.006 to +0.002) with identical segmentation quality scores (0.86 ± 0.11) as shown in Figure 2. T2f alone demonstrated robust performance (median DSC difference: –0.008, IQR: –0.016 to –0.001; segmentation quality: 0.76 ± 0.10), while other T2f combinations maintained excellent stability: T2f+T2w (median: –0.003, IQR: –0.011 to +0.001), T2f+T1n+T2w (median: –0.004, IQR: –0.009 to +0.001), and T2f+T1n (median: –0.006, IQR: –0.013 to +0.001). These combinations achieved segmentation quality scores of 0.77–0.78 ± 0.10, representing only a 0.08–0.09 decrease from baseline. Single-sequence models without T2f showed greater performance variability and larger systematic deviations: T1c alone (median: –0.049, IQR: – 0.079 to –0.027), T1n alone (median: –0.054, IQR: –0.087 to –0.033), and T2w alone (median: –0.029, IQR: –0.042 to –0.013). This pattern emphasizes T2f’s critical role in capturing complete tumor extent, including edematous components.

### Statistical Validation

Pairwise Wilcoxon signed-rank tests with Benjamini–Yekutieli false discovery rate correction revealed distinct statistical patterns that reinforce the performance findings across tumor regions (Figure 3).

For ET segmentation, all T1c-containing combinations showed non-significant differences from baseline, with T1c alone achieving p = 0.129 and T1c combinations demonstrating even higher p-values: T1c+T2f (p = 0.779), T1c+T1n (p = 0.788), and T1c+T2w (p = 0.822). These statistical results align directly with the minimal median DSC differences observed for these combinations (all ≤ 0.004). Conversely, all non-T1c combinations demonstrated highly significant performance degradation (p < 0.001), corresponding to their substantial median differences of –0.171 to –0.199.

For ED segmentation, the statistical analysis revealed that only T1c+T2f showed non-significant differences from baseline (p = 0.078), reflecting its superior median difference of –0.002 and highlighting the synergistic importance of combining both sequences. All other combinations, including T1c alone, were significantly inferior to the baseline (p < 0.001), demonstrating that neither sequence alone is sufficient for optimal edema delineation.

For WT segmentation, T1c+T2f achieved non-significant differences from baseline (p = 0.202) corresponding to its minimal median difference of –0.002, with T2f+T1n+T2w also approaching non-significance (p = 0.033). Single-sequence models without T2f were significantly inferior (p < 0.001), providing statistical confirmation of T2f’s essential role in whole tumor segmentation.

### Volumetric Accuracy Assessment

Volumetric evaluation showed distinct patterns across MRI sequence protocols, with important implications for reliable tumor measurement (Figure 4, Figure 5).

Protocols containing both T1c and T2f produced the most accurate segmentations for enhancing tumor (ET) and whole tumor (WT). The T1c+T2f combination achieved nearly perfect volumetric agreement with ground truth, with median differences close to zero in both ET (55 mm³, IQR: –788 to 1,279 mm³) and WT (–7 mm³, IQR: –3,069 to 3,664 mm³), and no significant difference from manual reference (adjusted p = 0.157 and p = 1.0). All T1c-containing protocols had non-significant ET errors (all adjusted p ≥ 0.114). In contrast, protocols lacking T1c—such as T2f, T1n, and T2w—showed pronounced over-segmentation in ET (e.g., T2f: –2,139 mm³, IQR: –7,723 to 790 mm³) and were all highly significant versus ground truth (all adjusted p < 0.001).

For WT, the highest accuracy was achieved by the baseline and T1c+T2f (both p = 1.0), as well as T2f+T2w (p = 0.255), T2f+T1n+T2w (p = 0.084), T1c+T1n+T2w (p = 0.075) and T2f (p = 0.075), which all yielded non-significant differences from ground truth (adjusted p ≥ 0.05). All other combinations returned statistically significant volumetric errors (adjusted p < 0.05).

Edema (ED) segmentation showed substantial over-segmentation and broad variability across all protocols. Although a few combinations—such as the baseline, T1c+T2f, T2f+T2w, and T2f+T1n+T2w—did not reach statistical significance, most protocols displayed significant errors, underscoring ongoing challenges for automated ED quantification.

In summary, T1c+T2f is the only simplified protocol consistently providing non-significant volumetric differences compared to ground truth for both ET and WT, enabling robust protocol reduction without accuracy loss in those compartments.

## Discussion

This study demonstrates that automated glioma segmentation can be delivered with clinical-grade accuracy using a streamlined two-sequence protocol (T1-contrast and T2-FLAIR), achieving non-significant volumetric differences relative to expert reference for enhancing and whole-tumor compartments and maintaining overlap-based segmentation performance comparable to a four-sequence reference, while reducing acquisition time. Critically, this identifies a minimal protocol that preserves volumetric and segmentation accuracy for targets most relevant to clinical decision-making, setting up the implications for response assessment and clinical implementation under contemporary practice standards, including RANO 2.0 (11).

Gliomas, particularly GBM, remain among the most difficult challenges in neuro-oncology due to their aggressive growth, infiltrative behavior, and marked heterogeneity on imaging, which complicate consistent measurement and longitudinal assessment in practice. Comprehensive overviews, including the MICCAI 2022 Brain Tumor Segmentation volume (32), underscore the central role of automated segmentation in both research and clinical neuro-oncology, and the need for models that are robust, generalizable, and clinically interpretable. Multicenter evaluations have demonstrated clinically acceptable glioblastoma segmentation across diverse datasets, and reviews emphasize that rigorous external validation and explicit management of heterogeneous clinical data—including missing sequences—are prerequisites for translation and reproducible response assessment (33,34). Nevertheless, many models still require complete, standardized MRI protocols, whereas real-world examinations frequently lack one or more sequences due to protocol variability, scan time constraints, motion, or technical limitations, limiting dependable deployment for quantitative response workflows (15).

This gap motivates development of tools like GlioMODA, explicitly designed to maintain performance under missing or heterogeneous input data. Such segmentation tools are prerequisites for reliable quantitative response assessment under RANO 2.0 (11). To this end, this study systematically evaluates segmentation and volumetric performance across 11 clinically relevant sequence combinations to define where accuracy is preserved and where it degrades when key inputs are omitted—information directly actionable for protocol design and site-level implementation. Addressing missing modalities during training, prior work has proposed sequence-dropout strategies to improve robustness when specific inputs are unavailable, without compromising accuracy when all modalities are present (35); these observations align with the current findings and support flexible frameworks for real-world adoption.

Precisely, our results demonstrate that GlioMODA achieves state-of-the-art segmentation performance across all tumor subregions, even when using fewer input sequences, and provide new insights into the sequence dependencies of automated glioma segmentation. T1-contrast enhancement (T1-CE) was indispensable for accurate enhancing-tumor delineation, models using T1-CE alone or in combination with other sequences performed comparably to the full-sequence reference, whereas omission of T1-CE produced systematic degradation. Conversely, T2-FLAIR was critical for capturing non-enhancing tumor extent, essential for accurate radiotherapy planning and clinical target volume (CTV) delineation (6), reinforcing its central role in whole-tumor assessment and longitudinal comparability. Consistently, Wilcoxon-test analysis revealed that T1-CE-containing combinations were not statistically inferior to the full protocol for enhancing tumor segmentation, while T2-FLAIR-containing combinations also maintained acceptable edema and whole tumor performance.

By evaluating panoptic quality (PQ), recognition quality (RQ), and segmentation quality (SQ), this analysis provides a comprehensive performance profile that couples lesion detectability with boundary precision. High RQ scores (>0.6) across all sequence combinations confirm robust tumor detection, while SQ more sensitively captures the effect of sequence completeness on boundary accuracy. This pairing—reliable recognition with context-dependent precision—supports longitudinal clinical workflows, maintaining dependable detection when boundary detail varies.

Our study demonstrates that a simplified MRI protocol using only T1-weighted contrast-enhanced (T1c) and T2-FLAIR (T2f) sequences yields volumetric measurements for enhancing and whole tumor regions that are nearly indistinguishable from expert manual ground truth, supporting streamlined acquisition with preservation of quantitative accuracy. This single key finding has strong clinical implications: it enables shorter scans and automated tumor volumetry that meets the performance requirements for current quantitative response workflows. The relevance of this result is underscored by the updated Response Assessment in Neuro-Oncology (RANO) 2.0 criteria, which formally permit volumetric response assessment alongside two-dimensional measures and emphasize standardized baseline and follow-up imaging (11). While RANO 2.0 retains two-dimensional assessments as the primary standard, it provides explicit volumetric thresholds—approximately 65% reduction for partial response and 40% increase for progression—that closely mirror traditional area-based cutoffs, thereby operationalizing quantitative categorization in clinical trials and care. Our data show that the T1c+T2f protocol achieves the reproducibility and precision needed for robust RANO 2.0 implementation, minimizing measurement error and enhancing the reliability of response categorization across timepoints and sites. Conversely, sequence combinations that omit either T1c or T2f are associated with systematic and often substantial volume errors that could misclassify response or progression under RANO 2.0 and compromise trial endpoints or treatment decisions. Importantly, the ability to maintain high volumetric accuracy with a simplified protocol addresses a central challenge highlighted in the RANO 2.0 update: ensuring standardized, longitudinally consistent imaging across diverse clinical settings with practical acquisition constraints. Taken together, these findings support automated, standardized MRI volumetry using the T1c+T2f protocol as a pragmatic pathway to RANO 2.0-aligned implementation in multicenter trials and routine practice. Although this study was limited to preoperative scans, the demonstrated protocol efficiency and compatibility with evolving standards provide a strong basis for prospective, longitudinal validation across treatment courses and institutions.

A recent study by De Sutter et al. systematically investigated the influence of different MRI modality combinations on automated glioblastoma segmentation, primarily using nnU-Net and SwinUNETR architectures (16). Consistent with our findings, they reported that T1-CE suffices for accurate enhancing-tumor and tumor-core delineation, and that T1-CE + T2-FLAIR achieves whole-tumor performance comparable to a four-sequence input; adding further modalities did not statistically improve accuracy and could introduce redundancy, although it reduced epistemic uncertainty. However, our study extends these findings in several important ways. First, we conducted a more comprehensive systematic evaluation by testing all clinically relevant sequence combinations, reflecting the spectrum of real-world protocol variability, rather than a subset, thereby directly informing protocol design under routine constraints. Second, we complemented overlap measures with recognition, segmentation, and panoptic quality (RQ/SQ/PQ) to separate lesion detectability from boundary accuracy, providing a more nuanced operational profile for clinical deployment. Third, we used Wilcoxon signed-rank tests and heatmap visualizations to rigorously determine which modality combinations are statistically non-inferior to the baseline, offering clearer guidance for protocol optimization. Finally, we release trained models and code as open source, supporting reproducibility, site-level validation, and PACS/DICOM integration—key prerequisites for clinical and multicenter trial implementation. Together, these advances emphasize technical rigor and practical implementability and deliver decision-ready guidance for implementation.

GlioMODA has been validated on the publicly available BraTS 2021 preoperative glioma dataset (1,251 training; 219 validation/test cases with expert segmentations), demonstrating robust performance across diverse imaging protocols. Recognizing that clinical integration of automated segmentation tools faces practical barriers including workflow adaptation, regulatory validation, and interoperability challenges, GlioMODA is provided as an open-source Python package available at https://github.com/BrainLesion/GlioMODA/ within the BrainLesion suite (36). This open release enables transparent, auditable use and local verification, and its modular design supports PACS/DICOM integration—providing a practical, validation-ready route to clinical deployment and multicenter trials, consistent with RANO 2.0 (11).

Future research directions include extending GlioMODA to pediatric gliomas (37) and rare tumor subtypes, where distinct imaging characteristics and age-specific biology may require adapted training strategies and evaluation endpoints, building on previous work investigating multiple pediatric brain tumor types (38). Prospective, multi-site validation in these populations will be essential to ensure safe clinical use and generalizability. Integration of advanced imaging modalities such as FET-PET represents a promising approach for enhanced molecular characterization. however, recent work shows both potential gains in volumetric accuracy and practical challenges, including volume underestimation and occasional segmentation errors (39), underscoring the need for careful calibration and cross-modality harmonization. The development of multitask models that simultaneously perform segmentation, tumor grading, and molecular prediction represents another promising direction, particularly where shared representations improve data efficiency and support end-to-end clinical workflows, as demonstrated by recent deep ensemble learning frameworks (40). Additionally, integration of GlioMODA into radiomics pipelines could enable personalized treatment strategies by linking imaging-derived features with molecular and clinical outcomes, supporting standardized, reproducible volumetry for response assessment consistent with contemporary criteria in neuro-oncology.

Several limitations must be acknowledged in our study. First, our analysis focused exclusively on preoperative glioblastoma cases from the BraTS 2021 dataset, which limits generalizability to postoperative imaging, other glioma subtypes, or pediatric populations where different imaging characteristics and segmentation challenges may apply. Second, while our systematic evaluation demonstrates robust performance across diverse sequence combinations, prospective validation in routine clinical workflows is required to confirm real-world applicability. Third, our current framework is optimized for structural MRI sequences and does not incorporate advanced imaging modalities such as perfusion MRI, diffusion tensor imaging, or molecular imaging techniques like FET-PET, which may provide additional diagnostic value in specific clinical scenarios. Fourth, the retrospective nature of our study using curated research datasets may underrepresent scanner/vendor variability, artifacts, and protocol deviations encountered in routine practice. Finally, while our volumetric accuracy analysis supports RANO 2.0 implementation, longitudinal, multi-timepoint validation across treatment response scenarios is needed to establish clinical utility for serial tumor monitoring and trial endpoints.

In conclusion, GlioMODA represents a practical advancement for automated glioma segmentation, demonstrating that T1-CE and T2-FLAIR sequences alone are sufficient for clinical-grade accuracy and are compatible with RANO 2.0. This approach enables workflow simplification, addresses key barriers to volumetric assessment, and provides a robust, open-source AI solution for routine neuro-oncology and future clinical trials.

## Data Availability

All data analyzed in this study are publicly available from the original sources. The Brain Tumor Segmentation (BraTS) 2021 dataset can be accessed at the official MICCAI portal (https://www.med.upenn.edu/cbica/brats2021/).
All code used for preprocessing, training, and evaluation is available at: https://github.com/BrainLesion/GlioMODA/.
No additional, non public human data were used.

https://github.com/BrainLesion/GlioMODA/

## Required Statements

### Ethics

This study used exclusively publicly available, de-identified BraTS 2021 data; accordingly, institutional review board approval and informed consent were not required.

### Authorship

Equal contribution: Julian Canisius and Josef Buchner contributed equally as co-first authors.

Shared senior authorship: Benedikt Wiestler and Florian Kofler contributed equally as co-senior authors.

### Data Availability

https://github.com/BrainLesion/GlioMODA/

## Supplementary Material

**Supplementary Figure S1.** Example of automatic segmentation by our T1c-only model (left column) and T2-FLAIR-only model (right column)

Axial views of the T1c and T2-FLAIR sequences are depicted in the upper and lower rows, respectively. The outline of the necrosis (in green), contrast-enhancing tumor (in red), and edema (in yellow) is shown. While the T1c-only model is able to precisely trace the outline of the contrast-enhancing tumor (upper left panel), it struggles with the delineation of the edema (lower left panel). The T2-FLAIR-only model behaves in the opposite way: the model does not delineate the contrast-enhancing tumor, while an accurate segmentation of the edema is achieved. The edema label on the opposite side is due to the tumor crossing the midline (not shown in this slice).

