## Supplementary figures and images for "GlioMODA: Robust Glioma Segmentation in Clinical Routine"

### Supplementary Figure S1.

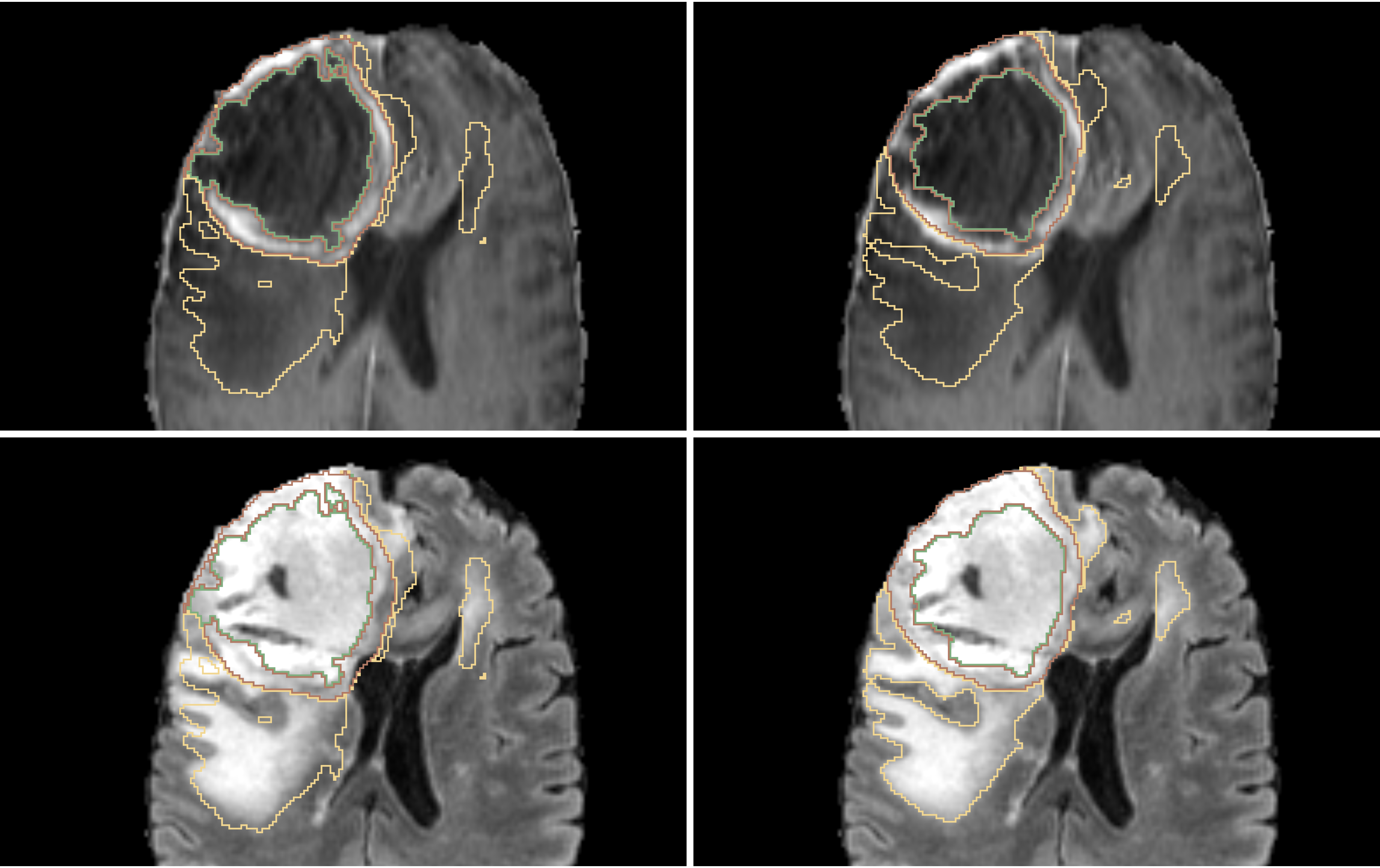
